# Bidirectional associations between PTSD severity and glycemic control in trauma-exposed women with type 2 diabetes

**DOI:** 10.1101/2025.11.26.25341097

**Authors:** Katharine J. Liang, Ronald G. Thomas, Kerry J. Ressler, Charles F. Gillespie, Rebecca C. Hendrickson

## Abstract

**Objective:** Posttraumatic stress disorder (PTSD) is linked with metabolic disturbance and increased risk of Type 2 diabetes (DM2), yet mechanisms explaining this connection have yet to be defined. Here we examine commonly hypothesized risk factors influencing the strength and directionality of the relationship between PTSD and DM2 severity in trauma-exposed black women with DM2.

**Methods:** We examined the relationships among PTSD severity (Clinician-Administered PTSD Scale, CAPS), glycemic control (A1c), age, smoking, body mass index (BMI), depression (Beck Depression Inventory, BDI) and medications in trauma-exposed Black women with DM2 recruited from an urban hospital between 2013-2015 (n=95) as a part of the Grady Trauma Project. Missing data were handled with multiple imputation. Relationships were assessed using lasso regression performed on each imputed dataset. Mediation analysis tested whether BMI or depression mediated associations between PTSD severity and glycemic control.

**Results:** Lasso regression identified BDI, ACE inhibitor/ARB/beta blocker use, and CAPS severity as predictors in the final model predicting A1c. In pooled linear regression analyses across imputed datasets, overall model fit was modest, and none of these predictors were statistically significant. Lasso regression in the reverse direction yielded BMI, BDI, ACE inhibitor/ARB use, and A1c for the final model predicting CAPS severity. In pooled analyses, model fit was substantially stronger and BDI emerged as the only statistically significant predictor (p<0.05). Mediation analyses indicated that BDI, but not BMI, significantly mediated the effect of A1c on CAPS severity, as well as to a lesser extent the reverse relationship, the effect of CAPS severity on A1c.

**Conclusion:** In this examination of the relationships between PTSD and DM2 severity, CAPS severity and A1c were found to be bidirectionally correlated in lasso regression models, with BDI emerging as a significant predictor and mediator of these relationships. BMI was not found to be a mediator in either direction. These results suggest the possibility of a reciprocal relationship between PTSD and DM2, where worsening of either condition may influence worsening of the other, where treating one condition in isolation may not be sufficient to prevent increased overall morbidity and mortality. Further research should include physiological interventions to ascertain causality in these relationships.

## Introduction

Individuals with posttraumatic stress disorder (PTSD) are at significantly increased risk of insulin resistance and associated cardiometabolic disorders. In studies of patients with PTSD, over half show evidence of metabolic dysfunction, with approximately 40% meeting criteria for metabolic syndrome (MetS) or prediabetes (PreDM)^1-4^ and an additional 10-20% meeting criteria for type 2 diabetes (DM2)^5,6^. A meta-analysis including 9 studies and over 9,000 subjects found that people with PTSD had almost doubled risk for comorbid MetS compared to controls without PTSD^3^. PTSD has also been found to double risk for both DM2 and cardiovascular disease^7-9^. Longitudinal studies additionally suggest that PTSD severity is prospectively associated with increased risk for both worsening MetS severity^10,11^ and developing new-onset DM2^12^, with PTSD symptoms severity predicting risk of DM2 in a dose-dependent fashion^13^. While mounting evidence suggests that trauma is linked to downstream disruption of both brain and metabolic processes, mechanisms explaining the connection between PTSD and metabolic dysfunction have yet to be fully elucidated.

Much of the research around drivers of IR has been focused on obesity; indeed there is a significant amount of evidence in the general population to support the metabolic syndrome model of abdominal obesity, hypertension, hyperlipidemia, and hyperglycemia as the pathophysiological precursor to DM2. A significant association between PTSD and obesity^14-16^ has led to speculation that, similar to known mechanisms of metabolic dysfunction, obesity may be the driver of the development of metabolic syndrome, IR, and progression to DM2 in PTSD.

Few studies have specifically examined the impact of obesity on the relationship between PTSD and DM2. However, two large longitudinal studies suggest that alternative mechanisms independent of obesity may contribute to the increased diabetes risk observed in this population. In the Nurses’ Health Study II, which included nearly 50,000 women, antidepressant use and BMI together accounted for roughly half of the association between PTSD and incident DM2^13^. Other health behaviors, including diet, exercise, and smoking, contributed minimally, leaving about half of the association unexplained. Similarly, the Millennium Cohort Study, involving nearly 45,000 US servicemembers, found baseline PTSD to be independently associated with diabetes risk even after adjusting for demographics and other mental and physical health conditions, including BMI^8^. In another mendelian randomization study, the relationship between PTSD and DM2 was only found to be partially mediated by obesity^17^.

Exploration of mental health conditions that may contribute to the relationship between PTSD and DM2 has revealed that depression may be a key factor in linking these conditions. In one large cohort study of people with and without PTSD, the increased risks of metabolic disorders and DM2 in the PTSD cohort were both attenuated when controlling for depression^18^. Another study in US Veterans found that the relationship between PTSD, obesity, and poor health behaviors was driven primarily through comorbid depression^19^. Furthermore, metabolic disturbances may play a role in the high prevalence of depression symptoms in PTSD. Over half of patients with PTSD have comorbid depression, with evidence from genetic risk factor and biological correlate studies suggesting that this trauma-related phenotype is distinct from isolated depression^20^. One potential biological explanation for this relationship is the connection between IR and depression. Several large meta-analyses have demonstrated an association between MetS^21,22^ or DM2^23^ and depression in the general population, with corroborating findings specifically in the context of PTSD^24^. This suggests the possibility that metabolic disturbances could also play a role in generating depressive symptoms or comorbid depression in PTSD. However, the majority of studies assessing the relationship between IR and unipolar depression are observational, limiting the ability to determine causality.

In an a priori review of the literature we had identified an increased risk of metabolic disturbances, including diabetes, in patients with PTSD. Some studies suggest that PTSD increases the risk of patients developing DM2. However, given that these diagnoses develop over long periods of time, there is limited evidence supporting directionality of these disorders or the nature of this observed connection. Given this question of directionality, we hypothesized that although statistical analyses cannot determine causality, that advanced statistical analyses may still provide some evidence to support the possibility of directionality with the data that is available. In previous analyses from a study of black women with DM2 and exposure to trauma, PTSD diagnosis was found to be associated with increased hemoglobin A1c^25^. In this secondary analysis, we further explore this relationship by factoring in the severity of PTSD in order to further tease out the strength of association in the relationship between PTSD (CAPS severity) and DM2 (A1c), as well as exploring the contribution of the following BMI, Beck Depression Inventory total score (BDI), age, lifetime smoking, and medications including antihyperglycemic medications (DM2rxtx), ACE inhibitors and angiotensin receptor blocking agents (ACEARBs), antidepressants, beta blockers, and HMG-CoA reductase inhibitors. Covariates included in the final models were selected based on previous studies linking each to the biological mechanisms of diabetes, PTSD, or both. The goal of our analyses was to further clarify relative contributions of each of these factors to the relationship between PTSD and DM2, information that could be important for informing treatment algorithms for patients in this difficult-to-treat population.

## Methods

In these analyses we relied on the power of advanced statistical analyses to suggest directionality from analyses conducted on an observational dataset. The use of continuous variables for our primary outcomes of interest, CAPS severity and A1c, allowed further power to test the magnitude of relationships. Furthermore, many studies investigating these relationships leave possible interacting factors such as medications, PTSD symptomatology, and comorbid psychiatric conditions unaccounted for. In the present analyses, lasso regression allowed inclusion of these potentially relevant variables in the pool for selection, providing an advantage of these analyses over many previous studies in this area. The combination of multiple imputation and lasso regression allowed us to take advantage of the relatively large and rich dataset that contained thousands of variables per participant to choose high quality predictor variables for imputation, while acknowledging that the need to impute missing data arose from the relatively small overall sample size of less than 100 participants. Significant filtering using well-established principles guiding multiple imputation variable selection was employed, as described above in methods. Lasso regression performed across 50 imputed datasets provided statistical stability and precision for pooled estimates.

### Participant population

This study is a secondary analysis of an observational dataset of trauma-exposed women with DM2, derived from a larger sample of black women with DM2 recruited from outpatient settings as a part of the Grady Trauma Project^26^. The subset of participants in this sample reported past exposure to trauma and were screened for exclusionary confounding factors as previously described^25,27^. Participants were not excluded for use of any medications, including diabetes or PTSD medications. Statistical analyses were conducted in R using the *psych* package, with missing data handled by multiple imputation using the *mice* package.

### Data analysis

The full dataset had 95 participants. In addition to the primary model variables of interest, hemoglobin A1c and PTSD severity as measured by the Clinician Administered PTSD Scale (CAPS severity), nine candidate model variables were selected for analysis a priori following manual review of all variables in the dataset, with selection guided by existing literature on PTSD and metabolic dysfunction to identify variables most relevant to hypothesized relationships. Missing data were handled using multiple imputation, and LASSO regression was applied across imputed datasets to identify the most relevant final model variables for inclusion in the final models as described below. Final analysis samples were extracted after imputation to include only those cases with observed, rather than imputed, primary endpoints (A1c and CAPS), resulting in a final analysis sample of 82 participants.

### Multiple imputation using mice

Multiple imputation was carried out for all primary and candidate model variables with missing values using the *mice* package in R^28^, with model variables defined as the target variables for multiple imputation. The number of missing values in the original dataset are shown in Supplemental Table 1. The original dataset contained 2914 potential predictor variables for selection. For each target variable, all primary and candidate model variables (except the target itself) were used for imputation, along with auxiliary predictor variables selected based on standard principles for selection of auxiliary variables for multiple imputation as previously described in detail^29^ and summarized here:

1. *High level filtering:* Potential predictor variables underwent initial filtering steps outlined in Supplemental Table 2.
2. *Ranking based on correlation and missingness:* To determine the strength of association with the target variable, Pearson correlations were calculated between each potential predictor and target variable. The variables were then ranked based on the correlation coefficient and the top 50 selected. To obtain adequate power for imputation, the proportion of non-missing values when the target variable was missing was also calculated for each of the top 50 potential predictor variables. Variables with >20% missingness when the target variable was missing were excluded. The correlation coefficient was then multiplied by the proportion of non-missing values for each potential predictor variable, generating a weighted predictor score that reflected both the strength of association and the amount of available data for cases where data is missing in the target variable.
3. *Final selection of predictor variables:* Final predictor variables consisted of the nine original target variables, with additional predictor variables selected from the top 50 predictors for each model variable based on weighted predictor score. Final selection favored predictors with higher weighted predictor scores. For each target variable, the final set of predictor variables was restricted to approximately one-third of the cases with complete data. To minimize multicollinearity in cases where multiple predictor variables were interrelated, for example in a family of predictors with one predictor variable reflecting the total score and the others reflecting its sub-components, only one in that family of predictors was chosen to represent the group.

Once custom auxiliary predictor matrices were defined for each target variable, multiple imputation was carried out using *mice*, creating 50 imputed datasets.

### LASSO Analysis

The Least Absolute Shrinkage and Selection Operator (LASSO) regression method^30^ was used to select which of the candidate model variables were included in each final model. Candidate model variables included CAPS severity, A1c, BMI, Beck Depression Inventory total score (BDI), age, lifetime smoking, and medications including antihyperglycemic medications (DM2rxtx), ACE inhibitors and angiotensin receptor blocking agents (ACEARBs), antidepressants, beta blockers, and HMG-CoA reductase inhibitors. Lasso regression was applied using the *glmnet* package in R, which allows for automatic standardization, with final coefficients returned on original measurement scales, preserving interpretability. To assess the most likely directionality of the relationships, lasso regression was applied in both directions, with separate analyses for each A1c or CAPS as the outcome variable. The threshold for inclusion in the final model was set at 50%, with candidate model variables selected in at least 25 of 50 rounds included in the final model.

### Mediation analysis

Mediation analysis was conducted using the potential outcomes framework to estimate Average Causal Mediation Effects (ACME), Average Direct Effects (ADE), and Total Effects. Bootstrap inference was employed with 1,000 simulations per imputed dataset.

## Results

### Lasso regression

Lasso regression examining A1c as the outcome variable resulted in four model variables selected in at least 25 of 50 imputations (Figure 1A): BDI, ACEARBs, Beta Blockers, and CAPS severity. Pooled analyses yielded a final model with Model R^2^ = 0.105, with none of the model variables selected demonstrating a statistically significant relationship with A1c (Table 1).

**Figure 1.**
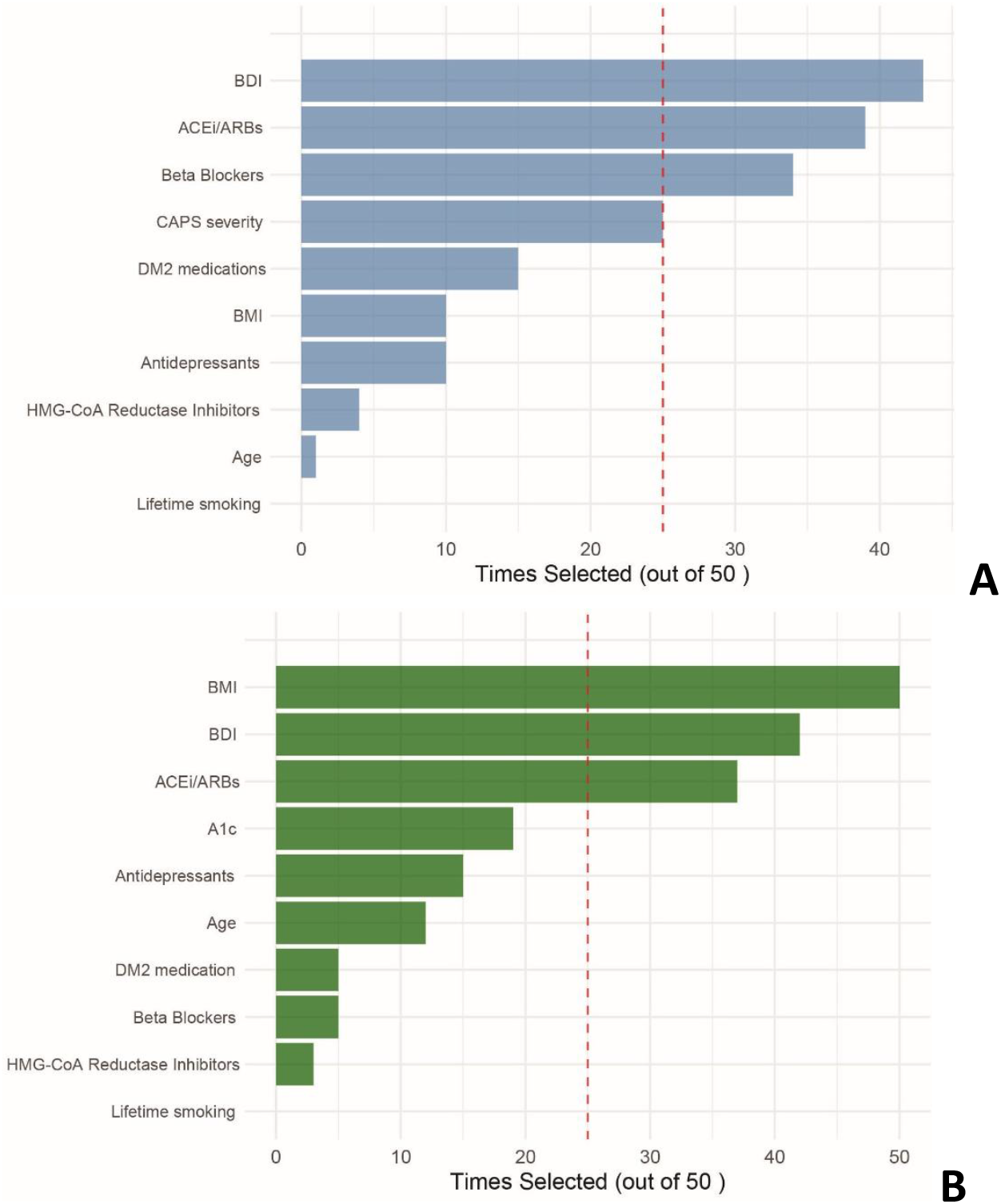
Lasso regression modeling variable selection for predictors of glycemic control as measured by A1c (A) and predictors of PTSD severity as measured by CAPS (B). Red line depicts a 50% threshold (25 of 50 imputations).

**Table 1.** Pooled results for lasso regression final model, with BDI, ACEi and ARBs, beta blockers, and CAPS severity as predictors of A1c. Model R^2^ = 0.105.

| Variable | Coefficient | SE | t value | p value |
| --- | --- | --- | --- | --- |
| BDI | 0.0141 | 0.0338 | 0.418 | 0.6763 |
| ACEi/ARBs | 0.0166 | 0.6009 | 0.028 | 0.9779 |
| Beta blockers | 0.4601 | 0.6607 | 0.696 | 0.4864 |
| CAPS_severity | 0.0454 | 0.0303 | 1.496 | 0.1346 |

Lasso regression examining CAPS as the outcome variable resulted in three model variables selected in at least 25 of 50 imputations (Figure 1B): BMI, BDI, and ACEARBs. In the lasso regression used to identify the strongest predictors of CAPS as the outcome variable, A1c did not meet the predetermined selection threshold; however, because the A1c-CAPS relationship was a prespecified primary hypothesis, we conducted additional sensitivity analyses to include A1c in the final model. Pooled analyses including A1c yielded a final model with R^2^ = 0.602, with BDI emerging as a statistically significant predictor of CAPS severity (p<0.05, Table 2). The final model including A1c as a predictor variable yielded similar patterns of association to the initial unpenalized analysis that did not include A1c (R^2^ = 0.592).

**Table 2.** Pooled results for lasso regression final model, with A1c, BMI, BDI, and ACE/ARB use as predictors of CAPS severity. Model R^2^ = 0.602.

| Variable | Coefficient | SE | t value | p value |
| --- | --- | --- | --- | --- |
| A1c | 0.5944 | 0.4154 | 1.431 | 0.1524 |
| BMI | -0.1368 | 0.1096 | -1.248 | 0.2120 |
| BDI | 0.8373 | 0.0848 | 9.879 | 0.0000 *** |
| ACEi/ARBs | -1.2563 | 2.1491 | -0.585 | 0.5589 |

### Mediation analysis

Mediation analyses were performed to test the hypotheses that BMI or BDI are mediators of the relationship between PTSD and DM2. There was a statistically significant indirect effect of A1c on CAPS through BDI (Table 3, ACME = 0.9373, p < 0.05), accounting for approximately 61.4% of the total effect. In the reverse direction there was also a statistically significant indirect effect of CAPS on A1c through BDI (Table 3, ACME = 0.0069, p < 0.05), although the magnitude was smaller, accounting for approximately 14.5% of the total effect. BMI was not a significant mediator of the relationship between CAPS and A1c in either direction.

**Table 3.** Summary of mediation analyses of testing BMI or BDI as mediators of the relationship between CAPS and A1c in either direction, indicating Average Causal Mediation (ACME), standard error (ACME_SE), Average Direct Effect (ADE), total effect (Total), and percent mediated.

| Scenario | ACME | ACME_SE | ADE | Total | % Mediated |
| --- | --- | --- | --- | --- | --- |
| 1: CAPS → BMI → a1c | 0.0000 | <0.01 | 0.0474 | 0.0475 | 0.1% |
| 2: a1c → BDI → CAPS | 0.9373 | <0.01 | 0.5904 | 1.5277 | 61.4% |
| 3: CAPS → BDI → a1c | 0.0069 | <0.01 | 0.0406 | 0.0475 | 14.5% |
| 4: a1c → BMI → CAPS | 0.0007 | <0.01 | 1.5270 | 1.5277 | 0% |

## Discussion

In this secondary analysis of data from trauma-exposed urban black women with DM2, we explored the relationships between PTSD severity and glycemic control using advanced statistical methods to infer directionality.

In these analyses we found that the strongest model explaining the relationships between PTSD and DM2 included A1c, BMI, BDI, and ACE inhibitor or ARB as predictors accounting for 60.4% of the variance in CAPS severity. The model in the reverse direction designating A1c as the dependent variable was much weaker, with BDI, ACEi and ARBs, beta blockers, and CAPS severity only accounting for 10.5% of the variance in A1c. These findings suggest a potential directionality in which worsening diabetes may be more strongly associated with worsening PTSD symptoms than the reverse, a pattern that contrasts with the directionality typically proposed in existing literature. While analyses from this single time point observational study dataset cannot determine causality, the asymmetry in the regression analyses suggest a directionality in this relationship that is worth exploring further in interventional longitudinal trials.

Obesity is one of the most studied drivers of DM2, with some suggestion in the literature that with the high prevalence of obesity and DM2 in PTSD, that the high prevalence of obesity in PTSD may be a driver of DM2 in this population. Given the conflicting evidence in the PTSD literature, we sought to test the hypothesis that obesity may be a mediating factor in the relationship between PTSD and DM2. Another driver of metabolic disturbance that has been implicated in psychiatric patients is depression. Though depression has gotten less attention than obesity, we also sought to use statistical mediation analysis to test the hypothesis that depression may be a mediating factor in these relationships.

In causal mediation analyses we found that the strongest mediating effect was that of BDI mediating the relationship between A1c and CAPS, with BDI accounting for 61.4% of the total effect. This result is consistent with previous findings that worsening metabolic disturbance is associated with depression and that similar relationships may play a role in explaining the connection between DM2 and PTSD. Importantly, BMI was not found to be mediator of the relationship between A1c and CAPS in either direction.

The lack of mediating effect of BMI on the relationship between PTSD and DM2 is perhaps surprising in a field where obesity is central to the canonical pathophysiology of DM2. One commonly proposed mechanism involves the role of obesity and sedentary lifestyle driving the connection between PTSD and IR, with multiple studies suggesting PTSD is associated with obesity and poor health behaviors^14,15,32^. However, this is in line with our hypotheses based on the limited literature available examining the role of obesity on metabolic health in this population, as well as our preliminary analyses in related datasets^31^. This finding is particularly noteworthy given that the standard of care for clinically addressing metabolic syndrome and prediabetes involves behavioral interventions addressing diet and exercise. Some studies demonstrate higher levels of physical activity and better diet in patients with PTSD^19,33^, raising the possibility that patients with severe PTSD may experience poor glycemic control despite a healthier diet and more active lifestyle. Furthermore, few studies have investigated the relative contribution of obesity and health behaviors to the relationship between PTSD and IR^7^, leaving gaps in our understanding of the mechanisms underlying these interrelationships. Our findings raise the possibility that obesity may contribute less to diabetes progression in PTSD than previously thought, a question that warrants further investigation.

Instead, the mediating effect of depression symptoms on the relationships between PTSD and glycemic control, while also limited by a low level of evidence, is an effect that we observed in this study in both directions and is consistent with previous literature in this area. These analyses build on previous analyses of this dataset, in which PTSD, but not MDD diagnosis, was found to be associated with increased A1c^25^. Taken together, these findings together suggest that it is possible that depression symptoms in the context of PTSD may have a different influence on glycemic control than depression when examined in isolation.

A complicated diagnostic nuance is the distinction between depressive symptoms and depression as an independent primary diagnosis, such as in the context of major depressive disorder (MDD). Given that one of the 4 clusters of PTSD symptoms specified in DSM5 is focused on negative affect and cognitions that are strongly overlapping with depression for diagnosis, in this study we conceptualized depression as measured by BDI as a component of PTSD symptom expression. While it is possible that some participants in our sample meet criteria for comorbid PTSD and MDD, this was not examined in present analyses.

This analysis has several limitations. First, as an observational dataset, this analysis cannot determine causality. However, the findings in this study may serve to inform design of future interventional studies to determine causal relationships. Secondly, the population of participants with preexisting DM2 and the nuance of the outcome measures tested in this study may explain the difference in our results compared to previous findings in the literature. In this study, participants were recruited who had preexisting DM2, and the outcome variable used to estimate worsening diabetes was A1c, a measure of glycemic control. This is different from previous studies that examined patients with PTSD and looked at risk of developing metabolic syndrome, prediabetes, or DM2 over time. It is possible, for example, that PTSD symptoms may drive risk of developing new onset DM2, and that once patients develop DM2, that worsened glycemic control feeds back on worsening PTSD symptoms. There is also the possibility of poor adherence to diabetes medications as a factor that we did not control in this analysis that should be assessed in future studies. Further studies examining the relationships between PTSD and metabolic disturbances at various stages of disease progression will be necessary to tease out the nuances of the interconnections between these disorders that may evolve at different stages of disease progression.

Many of the measures we used to estimate worsening disease, such as A1c as an approximation of worsening diabetes and BMI as an indicator of obesity, are indirect measures that do not fully reflect the severity of those respective conditions. To more accurately study these conditions, future studies should employ more sensitive and specific measures to estimate the severity of disease, such as using a glucose tolerance test as a measure of insulin sensitivity or a combination of more sensitive biometric measures such as body composition analysis as a measure of obesity.

In summary, our findings suggest a bidirectional relationship between PTSD and diabetes, with predictors explaining a greater proportion of variance in CAPS severity than A1c. Depression symptoms significantly mediated the relationship between CAPS severity and A1c in both directions, with depression playing a more substantial mediating role for the pathway from A1c to CAPS than from CAPS to A1c. Further studies are warranted to determine the nature and extent of these relationships.

## Supplemental materials

**Supplemental Table 1.** Full dataset, binary variables.

|  | Full dataset |  |  |  | After excluding participants with incomplete primary endpoints |  |  |  |  |
| --- | --- | --- | --- | --- | --- | --- | --- | --- | --- |
| **Participant responses** | **0** | **1** | **NA** | **Complete cases** | **0** | **1** | **NA** | **Complete cases** | **# of predictor variables used in imputation** |
| DM2rxtx | 7 | 66 | 22 | 24 | 7 | 61 | 14 | 68 | 22 |
| ACEARBs | 21 | 49 | 25 | 23 | 20 | 45 | 17 | 65 | 21 |
| Antidepressants | 56 | 15 | 24 | 23 | 53 | 13 | 16 | 66 | 22 |
| BetaBlockers | 55 | 16 | 24 | 23 | 51 | 15 | 16 | 66 | 22 |
| HMG CoA Reductase Inhibitors | 32 | 40 | 23 | 24 | 32 | 35 | 15 | 67 | 22 |
| Smoking lifetime | 19 | 36 | 40 | 18 | 17 | 31 | 34 | 48 | 16 |

**Supplemental Table 2.** High level filtering for predictor variable selection.

| Filter for predictors | n remaining after filter applied |
| --- | --- |
| Step 1: Removed non-numeric variables | 2463 |
| Step 2: Removed variables with zero and near-zero variance | 1843 |
| Step 3: Removed variables with $\geq 30\%$ missingness | 1258 |

## Data Availability

All data produced in the present study are available upon reasonable request to the authors

